# Statistical analysis plan for the feasibility trial assessing external trigeminal nerve stimulation versus sham stimulation for ADHD in children and adolescents aged 7–17years (the eTNS4ADHD trial)

**DOI:** 10.64898/2026.10.30.26365900

**Authors:** Sophie Juul, Markus Harboe Olsen, Sabrina Katja Nestved, Maria Quistgaard, Henriette Edemann-Callesen, Christel-Mie Lykke Huus, Caroline Karstoft, Ingrid Barisic, Elin Bjarnadottir, Aida Bikic, Pia Jeppesen, Ørjan G Martinsen, Fred Johan Pettersen, Jane Lindschou, Christian Gluud, Thorsten Schumann, Janne Walløe Vilmar, Ole Jakob Storebø

## Abstract

**Background:** External trigeminal nerve stimulation (eTNS) is a non-invasive intervention being investigated as a potential treatment for attention-deficit/hyperactivity disorder (ADHD) in children and adolescents. The eTNS4ADHD trial aims to assess the feasibility of conducting a larger randomised clinical trial comparing active eTNS with sham stimulation. This paper presents the prespecified statistical analysis plan for the trial.

**Methods:** The eTNS4ADHD trial is a multicentre, parallel-group, randomised pilot and feasibility trial involving children and adolescents aged 7–17 years with ADHD. Participants are allocated to four weeks of active eTNS or sham stimulation. Feasibility outcomes include recruitment, treatment compliance, acceptability, follow-up completion, and concomitant treatment use. Exploratory clinical outcomes include ADHD symptoms, school absence, emotional lability, functional impairment, quality of life, sleep quality, and adverse events. All analyses will follow the intention-to-treat principle. Feasibility outcomes will primarily be assessed using proportions with 95% confidence intervals, while exploratory clinical outcomes will be analysed using regression models or non-parametric methods, as appropriate. Two independent statisticians blinded to treatment allocation will conduct the analyses, and blinded conclusions will be prepared before unblinding.

**Discussion:** This statistical analysis plan was finalised before unblinding to promote transparency and minimise the risk of selective analyses and reporting. Given the feasibility design and limited statistical power, exploratory clinical findings will be considered hypothesis-generating rather than confirmatory. The results will inform the feasibility, outcome selection, sample size estimation, and statistical methodology of a future definitive randomised clinical trial evaluating eTNS for ADHD in children and adolescents.

**Trial registration:** ClinicalTrials.gov, NCT06655610.

## Introduction

This statistical analysis plan describes the pre-planned statistical analyses for the feasibility trial assessing external trigeminal nerve stimulation versus sham stimulation for ADHD in children and adolescents aged 7–17 years (the eTNS4ADHD trial).

The design of the eTNS4ADHD trial has been described in detail previously [1]. In short, the trial is a multi-centre, parallel group, investigator-initiated, randomised clinical pilot and feasibility trial. The eTNS4ADHD trial was pre-registered at ClinicalTrials.gov (identifier: NCT06655610), and carried out in compliance with the Declaration of Helsinki [2]. The trial was approved by the Danish Medical Ethics Committee (approval number: 2401733), and by the Regional Data Protection Agency (approval number: P-2024-16214). This statistical analysis plan was finalised and approved by the steering committee before unblinding of trial data.

### Outcomes

**Table 1** below presents an overview of the outcomes, measures, data types, and assessment schedule stated in the eTNS4ADHD protocol.

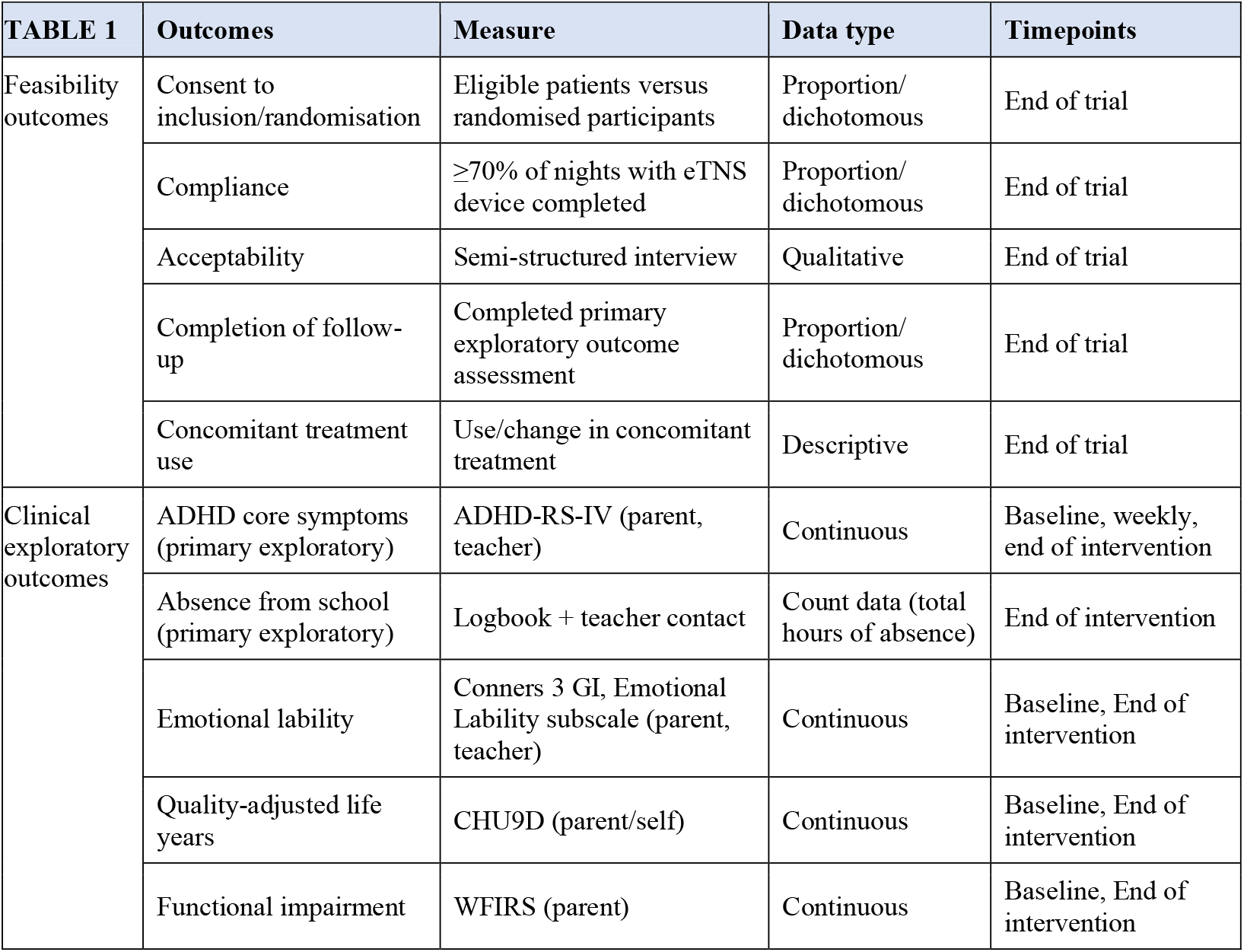
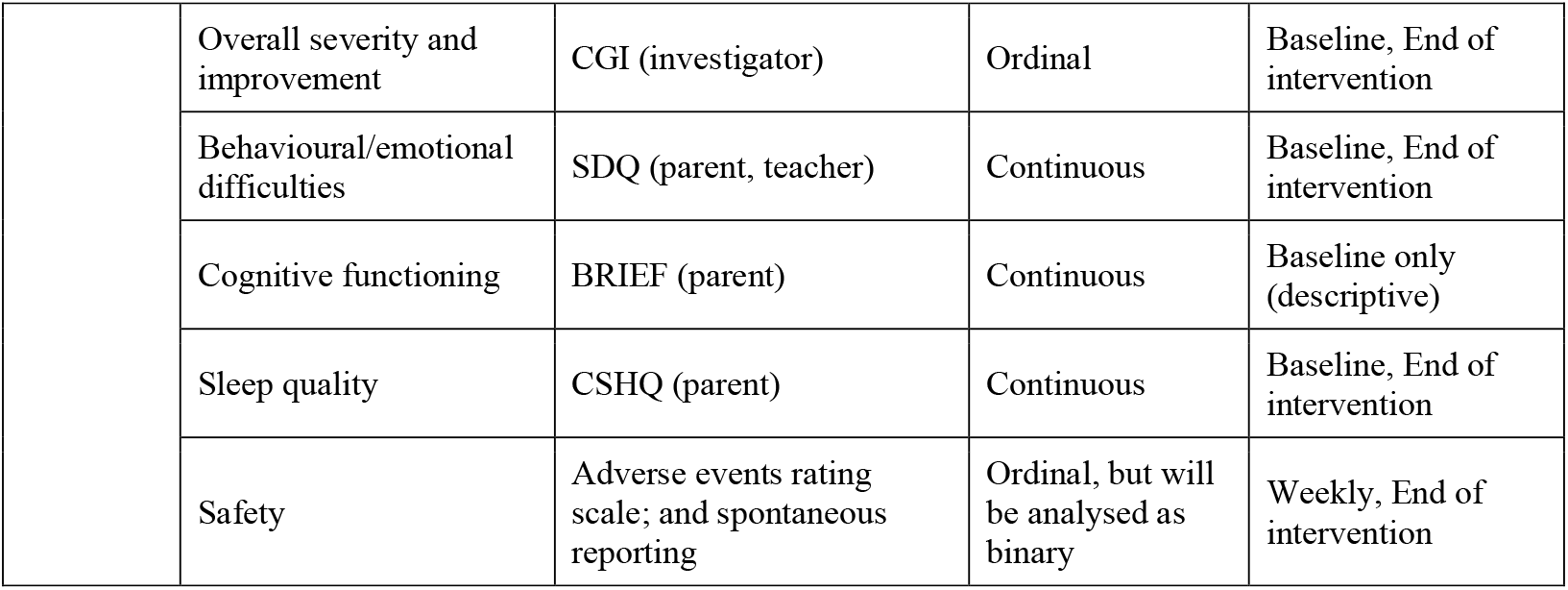

**Table 2.** Baseline characteristics (simulated data)

|  | Active eTNS | Sham | Overall |
| --- | --- | --- | --- |
| **n** | 30 | 30 | 60 |
| Age, years (mean (SD)) | 11.4 (2.5) | 11.0 (2.7) | 11.2 (2.6) |
| **Sex (%)** |  |  |  |
| Male | 20 (66.7) | 19 (63.3) | 39 (65.0) |
| Female | 10 (33.3) | 11 (36.7) | 21 (35.0) |
| **ADHD presentation, DSM-5 (%)** |  |  |  |
| Combined | 18 (60.0) | 17 (56.7) | 35 (58.3) |
| Predominantly inattentive | 9 (30.0) | 10 (33.3) | 19 (31.7) |
| Predominantly hyperactive-impulsive | 3 (10.0) | 3 (10.0) | 6 (10.0) |
| **Medication status at entry (%)** |  |  |  |
| Treatment-naïve | 12 (40.0) | 12 (40.0) | 24 (40.0) |
| Previous stimulant use, not current | 6 (20.0) | 6 (20.0) | 12 (20.0) |
| Current stable stimulant medication | 12 (40.0) | 12 (40.0) | 24 (40.0) |
| **Trial site (%)** |  |  |  |
| Region Zealand | 10 (33.3) | 11 (36.7) | 21 (35.0) |
| Region of Southern Denmark, site 1 | 12 (40.0) | 11 (36.7) | 23 (38.3) |
| Region of Southern Denmark, site 2 | 8 (26.7) | 8 (26.7) | 16 (26.7) |
| Region of Southern Denmark, site 3 | 7 (26.7) | 6 (26.7) | 10 (26.7) |
| IQ, WISC-V (mean (SD)) | 98.5 (12.1) | 99.2 (11.4) | 98.8 (11.7) |
| ADHD-RS-IV total score, baseline (mean (SD)) | 38.4 (7.2) | 37.9 (6.8) | 38.1 (7.0) |
| CSHQ total score, baseline (mean (SD)) | 52.3 (8.9) | 51.6 (9.4) | 51.9 (9.1) |

Missing data: IQ, 1.7%. Values are illustrative/simulated and do not represent real participants.

**Table 3.** Feasibility outcomes.

| Outcome | Protocol target (95% CI) | Simulated result, n/N (%) | Simulated 95% CI | Feasible? |
| --- | --- | --- | --- | --- |
| Consent to inclusion/randomisation | >50% (40.2–59.6%) | 66/90 (73.3%) | 63.6–83.0% | Yes |
| Compliance with treatment | ≥70% (58.4–81.6%) | 46/60 (76.7%) | 65.2–88.2% | Yes |
| Completion of follow-up | ≥90% (82.4–97.6%) | 55/60 (91.7%) | 83.8–99.5% | Yes |
| Acceptability of the intervention | Assessed qualitatively (no numeric threshold) | n/a – thematic analysis | n/a | n/a |

Green shading indicates the simulated result meets the protocol-specified feasibility threshold. Values are illustrative/simulated.

**Table 4.**
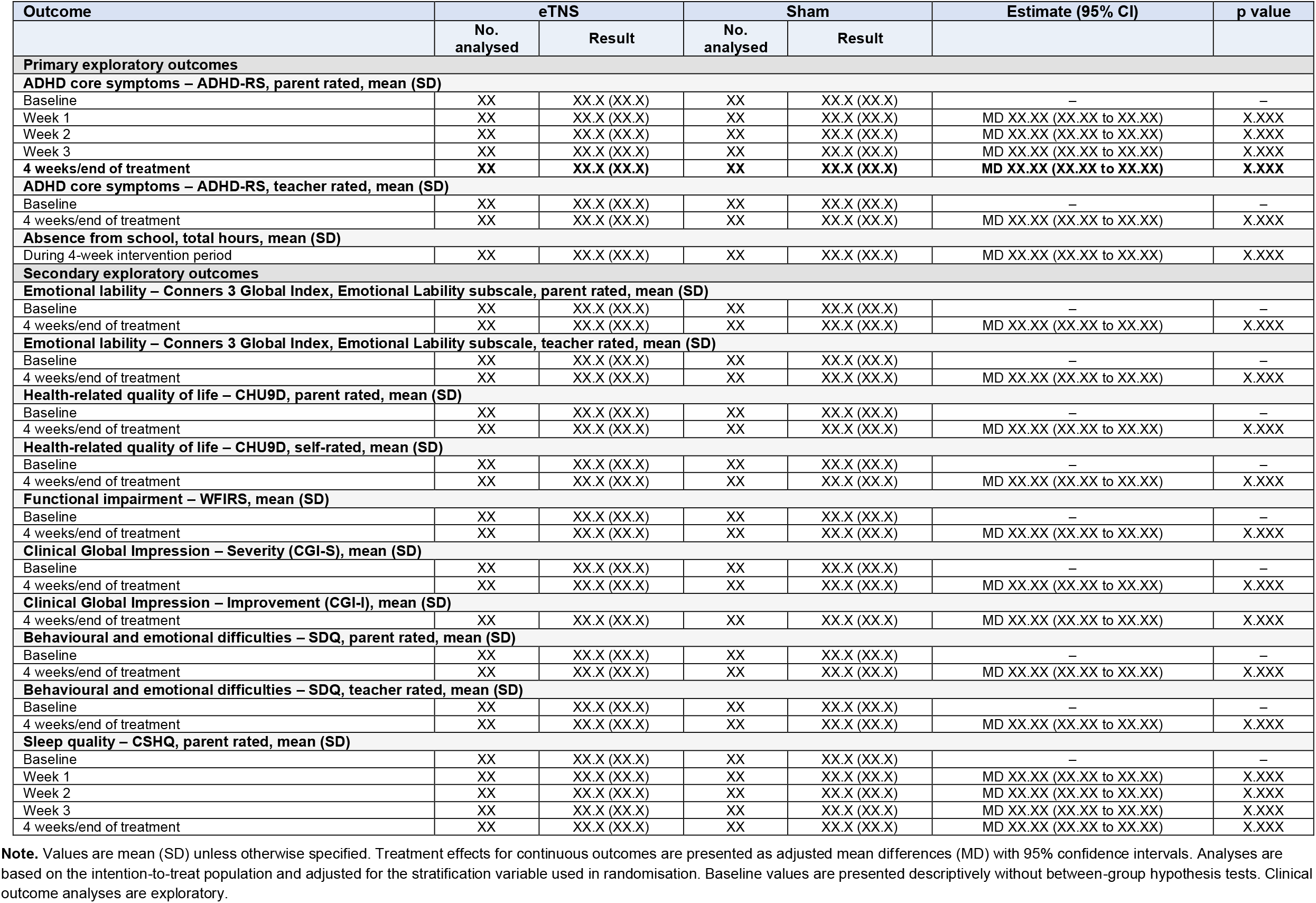
Exploratory clinical outcomes (see supplementary material) Exploratory clinical outcomes in the intention-to-treat population

### General analysis principles

All analyses will be performed on an intention-to-treat basis. All randomised participants will be analysed in the group to which they were allocated, irrespective of the amount of treatment received. Baseline characteristics will be reported descriptively by group (mean (SD) or median (IQR) for continuous variables; n (%) for categorical variables), with no between-group testing at baseline given the randomised design.

All regression analyses of exploratory clinical outcomes will be adjusted for the stratification variable used at randomisation, trial site: Region South or Region Zealand.)

Given the exploratory nature of the clinical outcomes and the trial not being powered for hypothesis testing (e.g. 48% power for the primary exploratory outcome), a significance threshold of p<0.05 will be used for descriptive/hypothesis-generating purposes only, with no correction for multiple comparisons. All exploratory results will be interpreted with caution and as signals to inform outcome selection, sample size, and design of a future definitive trial, not as evidence of efficacy or safety.

Two statisticians, blinded to group allocation (‘A’ and ‘B’), will independently perform the analyses according to this statistical analysis plan and each will produce an independent statistical report. Any discrepancies will be discussed and resolved before unblinding. The agreed statistical report will be version-controlled. Two blinded conclusions will be drafted by the steering group prior to unblinding (one assuming A = active eTNS, one assuming A = sham); once unblinding occurs, the correct conclusion will be retained without revision.

Statistical analyses will be performed in R and Stata.

### Analysis of feasibility outcomes

Feasibility outcomes are measured at the end of the 4-week treatment period and will be presented as proportions with 95% confidence intervals (CI), calculated using a 1-sample proportions test with continuity correction, with the confidence limit adjusted to a maximum of 100% (in line with the thresholds prespecified in the protocol). The lower bound of the 95% CI will be used as the conservative indicator of feasibility for progression to a full-scale trial, consistent with the protocol’s prespecified criteria.

#### 1) Consent to inclusion and randomisation

The number of eligible participants who consent and are randomised will be compared with the total number of participants assessed for eligibility. A difference (randomised/eligible) above 50% (protocol-specified 95% CI 40.2–59.6%) will be considered feasible. We will use the lower limit of the confidence interval as the threshold for feasibility.

#### 2) Compliance with the intervention

Compliance will be calculated separately for the active and sham groups as the proportion of participants completing treatment on ≥70% of the 4-week nights (protocol-specified 95% CI 57– 80%), where one night requires ≥5 hours of uninterrupted stimulation and an abruption of <4 consecutive nights is permitted. Compliance data will be derived from the nightly logbooks and presented by group. We will use the lower limit of the confidence interval as the threshold for feasibility.

#### 3) Acceptability of the intervention

Acceptability will be assessed qualitatively via a semi-structured interview guide (four pre-defined questions) and analysed and reported narratively/thematically outside the quantitative statistical report; no formal statistical testing will be applied to this outcome.

#### 4) Completion of follow-up

Defined as completion of the primary exploratory outcome assessment (ADHD-RS-IV) at end of treatment. The number of participants completing the assessment will be compared with the total number randomised. A completion rate ≥90% (protocol-specified 95% CI 82.4–97.6%) will be considered acceptable for progression to a full-scale trial. We will use the lower limit of the confidence interval as the threshold for feasibility.

#### 5) Use of concomitant treatment

Concomitant treatment/medication use and any changes during the trial will be summarised descriptively (n, %) for each group at end of trial; no formal hypothesis test is planned given the exploratory nature of this outcome.

### Analysis of exploratory clinical outcomes

Exploratory clinical outcomes will be analysed using the same general approach for a potential future full-scale trial, selecting the analysis method appropriate to each outcome’s distribution and assessment schedule. All results will be presented as effect estimates (mean difference, risk ratio, or median difference as appropriate) with 95% CIs (or 95% Hodges-Lehmann confidence interval), and interpreted descriptively given the limited power of this feasibility trial. Given the low number of participants an univariable analysis (Fisher’s test, Student’s t-test, or Wilcoxon rank sum test, as appropriate) will be carried out as sensitivity analysis.

#### Continuous outcomes assessed at baseline and end of treatment only

Continuous outcomes will be analysed using linear regression of the end-of-treatment score, adjusted for the corresponding baseline score and the stratification variable: trial site. Results presented as adjusted mean differences (active vs. sham) with 95% CI.

#### Continuous outcomes with weekly repeated measures (ADHD-RS-IV parent-rated, CSHQ parent- and teacher-rated)

The primary between-group comparison will use the end-of-treatment assessment, analysed by linear regression adjusted for baseline score, stratification variable, and site, as above. Weekly trajectories will additionally be presented graphically by group for descriptive purposes only; given the feasibility trial’s limited power and the risk of inflating type I error across multiple weekly comparisons, no formal repeated-measures hypothesis test is planned as a primary analysis. As an exploratory analysis, we will perform a mixed-effects model for repeated measures (MMRM) with fixed effects for treatment, time, treatment-by-time interaction, baseline score, and stratification variable (site).

#### Ordinal outcome (CGI)

Presented descriptively (n, % per category, or median/IQR) by group at end of treatment. If a between-group comparison is desired, this can be analysed using ordinal logistic regression or, if proportional-odds assumptions are not met, dichotomised and analysed by logistic regression.

#### Count data outcomes (absence from school – total hours)

Presented as medians and interquartile ranges per group and analysed using the Mann-Whitney U test; Hodges–Lehmann estimates of the location shift with 95% CI will be presented to quantify the between-group difference.

### Safety

Adverse events, recorded via the adverse events rating scale weekly, at end of treatment and via any spontaneous reports during the trial, will be summarised descriptively by group: number and proportion of participants reporting at least one adverse event, and number/type/severity of individual events. Height, weight, and vital signs at baseline and end of treatment will be summarised descriptively by group (mean (SD) or median (IQR), and change from baseline). Given the small sample size, formal between-group statistical testing of adverse events will be limited to descriptive comparison (proportions with 95% CI); Fisher’s exact test may be used for sparse cell counts if a comparison is presented. In such case, the adverse events data will be analysed as dichotomous outcome (participants with at least one SAE versus participants with no SAE in both groups).

### Handling of missing data

No imputation method (e.g. multiple imputation) will be used to handle missing data in this feasibility trial. The extent and pattern of missing data will instead be reported in detail (n and % missing per outcome, per group, per timepoint) in the statistical report, to inform data-collection procedures and missingness assumptions for the design of the future full-scale trial.

### Assessment of underlying statistical assumptions

Assumptions of the linear regression models (normality and homogeneity of residuals) will be assessed visually using histograms and quantile-quantile plots of residuals, and residuals-versus-fitted-value plots; logarithmic transformation or robust standard errors will be considered if deviations are identified. For logistic regression models, overdispersion will be assessed (deviance/degrees of freedom), and Fisher’s exact test will be used instead where event counts are low (substantially below the rule-of-thumb of 10 events per variable). Given the small sample size of this feasibility trial, confidence intervals and p-values should be interpreted with caution regardless of assumption checks.

Additionally, trial site may be omitted as an adjustment variable, if assumptions are not met.

### Subgroup analyses

No subgroup analyses are planned in this feasibility trial. The stratification variable (trial site) will be included only as an adjustment covariate. Subgroup analyses are deferred to the future, larger randomised trial.

### Statistical reports and version control

A statistical report will be developed and version-controlled prior to the randomisation of the last participant, in line with the protocol. Following completion of the trial, two statisticians blinded to allocation will independently analyse the data and produce reports labelled ‘A’ and ‘B’; a third party will compare these reports and resolve discrepancies before unblinding. The final statistical report and both independently produced blinded reports will be retained as supplementary trial documentation.

### Illustrative tables based on simulated data

The tables below illustrate the planned format of the statistical report using simulated data. Values are entirely fictitious and are included only to demonstrate table structure, layout, and the level of detail expected in the final report; they must be replaced with real trial data - including correct missing-data annotations - once available.

## Data Availability

Data availability is not applicable to this design/protocol paper

## References

1. Edemann-Callesen H, Huus C-ML, Karstoft C, Bjarnadóttir E, Bikic A, Jeppesen P, et al. External trigeminal nerve stimulation versus sham stimulation for attention deficit hyperactivity disorder in children and adolescents aged 7–17 years: study protocol for a pilot and feasibility randomized clinical trial. European Child & Adolescent Psychiatry. 2025;34(12):3833–42.

2. World Medical Association. World Medical Association Declaration of Helsinki: ethical principles for medical research involving human subjects. Jama. 2013;310(20):2191–4.

